# The effect of pulsatile Versus nonpulsatile cardiopulmonary bypass on renal function in patients undergoing coronary artery bypass graft surgery, a randomized controlled trial

**DOI:** 10.1101/2023.02.07.23285629

**Authors:** Saeed Khademi, Marzieh Zamani Jahromi, Mohammad Ghazinoor, Amirmohammad Farrokhi, Mohamad Hosein Bahmani Kazerooni, Masoud Najafi

## Abstract

**Background:** Coronary artery bypass grafting is of the most major surgeries performed around the world. Even though advances are achieved in the surgical technique, a relatively high complication rate regarding circulation is still observed. These complications are believed to be related to cardiopulmonary bypass flow types, pulsatile and nonpulsatile. With renal complications being one of the most important ones, we aim to evaluate the effect of choice of these two flow types on patients’ renal function in a randomized controlled trial.

**Method:** The study is a double blind randomized clinical trial. Patients with left ventricular dysfunction who were candidates for CABG and were between the ages of 40 to 75 were included in this study. The patients then were randomly assigned into two groups of intraoperative pulsatile and nonpulsatile flow type. The patients renal function markers such as 24-hour urine output, blood urea nitrogen and serum creatinine levels and creatinine clearance were evaluated before and CABG and afterwards in the ICU ward. The results were then analyzed using SPSS 23 software.

**Results:** of the initial 80 patients enrolled in this study, 16 patients were dropped due to unwillingness to continue follow-up and limitation of data gathering. Patients demographic data between two groups did not differ significantly. No statistically significant difference was observed between the 24 patients undergoing surgery with pulsatile flow and 40 with nonpulsatile flow regarding renal function. Both groups had a decrease in creatinine clearance during their ICU stay. Patients in the pulsatile flow group had less intubation time, less need for blood transfusion but more bleeding after the surgery.

**Conclusion:** Our study indicated that there is no difference between the use of pulsatile versus nonpulsatile flow regarding patients’ renal outcome. Our participants had a relatively broader age range than similar studies, including younger patients. This plus having an acceptable number of patients evaluated may illustrate that the differences in these two flow types may be dependent on other risk factors depending on the studied population. Further investigations with focal groups could lead us towards a better understanding how these two flow types differ.

## Background

Coronary artery bypass grafting (CABG) is one of the most common major surgeries performed worldwide, 400,000 being performed annually(1). This procedure is performed with the aid of cardiopulmonary bypass(CPB) as the artificial circulatory device(2). Despite advances in surgical techniques, a systemic perfusion or microcirculatory complication rate of up to 40% is observed in patients undergoing these operations(3, 4). Complications vary from sepsis and thrombosis to kidney damage which could lead to renal failure, gastrointestinal bleedings, pulmonary injury and neurological dysfunction added to longer hospitalization and need for multiple surgeries which contribute to increased costs(3-5). These complications are noted to probably be due to different flow types of CPB, pulsatile and non-pulsatile(6). Despite upgrades and improvements in technology in CPB, it is yet considered a non-physiological scenario(7). Pulsatile flow, though lacking a universal definition, is broadly accepted as when pulse pressure is greater than 15 mmHg and a pulse pressure beneath that counts as non-pulsatile(8).

Pulsatile flow may be preferred since it resembles the physiological state more, increases endothelial shear stress leading to synthesis of endothelial vasodilators and does not lead to capillary collapse, which is believed to induce tissue hypo-perfusion(9). Furthermore, evidence has showed that non-pulsatile flow leads to higher levels of sympathetic activity, contributing to a poor outcome and disease progression(10).

Even though findings in several studies point to a general preference towards use of pulsatile flow during CPB, the evidence have not been conclusive for a definitive superiority due to the results not being unanimous in numerous trials(11, 12). This could be due to different study protocols and small sample sizes in each study or lack of having a definitive protocol(13). In this randomized controlled trial, we aim to compare the renal outcome of patients undergoing coronary artery bypass graft surgery (CABG) using CPB in pulsatile and non-pulsatile flow groups.

## Method

### Study design

This study is a double-blind, randomized controlled trial (RCT). The inclusion criteria were patients between the ages of 40 to 75 years old who had left ventricle dysfunction defined by an ejection fraction (EF) of less than 40% whom were candidates for elective CABG in Namazi hospital, Shiraz. Patients who had an anesthesia risk class of over 3 based on American society of anesthesiologists classification system, systemic infection or sepsis, liver dysfunction, prior history of stroke, autoimmune disease, simultaneous need for CABG and valve replacement, previous CABG, history of immunosuppressant therapy, chemotherapy or radiotherapy, serum creatinine of over 1.5mg/dl and those who weren’t willing to participate in the study were excluded from our study.

### Intervention

Patients were allocated into two groups of pulsatile and non-pulsatile flow in CPB by using a random number generator by a researcher. All patients received a general anesthesia dosage of 0.1 to 0.2 mg/kg midazolam, 0.3 to 1 µg/kg sufentanil, 1 to 2 mg/kg sodium thiopental, and 0.1 to 0.2 mg/kg pancuronium bromide. After intubation, anesthesia maintenance was performed via intravenous drugs such as propofol. Morphine was infused with a dose of 0.1mg/kg if indicated. When CPB was established with a cardiac index of 2 to 2.4 Lit/min/m^2^, mean arterial pressure (MAP) between a range of 50 to 90 mmHg was established. Positive inotropic drugs such as epinephrine and norepinephrine were used to maintain the MAP. In the case of lack of urine output, 0.5 to 1 mg/kg IV furosemide was administered and recorded both intraoperative and after operation in the intensive care unit (ICU) ward.

Following sternotomy and dissection of proper graft, cannulation was performed and CPB was established. At this stage, flow types were applied to CPB according to prior allocation of patients. After clamping of the aorta, a cardioplegic solution consisting of 25mEq sodium bicarbonate, 30 to 40mEq potassium chloride, 40 mg lidocaine, 1g magnesium sulfate in one liter of normal saline solution was injected via the cardioplegic cannula. Time of asystole phase was recorded and then distal anastomoses of venous grafts and coronary arteries is performed while maintaining a body temperature of 34° Celsius. Afterwards, gradual increase of body temperature is initiated and simultaneous proximal anastomoses is performed. After chest tube insertion and evaluation for bleeding and proper anastomosis, patient is disconnected from the CPB device and then transferred to ICU ward for mechanical ventilation and cardiac monitoring. The patient then is transferred to ward if no complication occurs. All operations were performed by one surgeon to avoid the possible surgeon related bias.

### Clinical and laboratory monitoring

Baseline laboratory data was gathered regarding renal function such as blood urea nitrogen (BUN) and serum creatinine levels one day prior to operation. BUN, serum creatinine, 24-hour urine output was evaluated daily as renal function markers in addition to intraoperative urine output. Central venous pressure (CVP) was regulated to be in the range of 8 to 12 cmH_2_O to eliminate intravascular volume effects on BUN. Liver and cardiac enzyme tests were carried out before and after surgery.

### Endpoints

The primary endpoint is the difference of renal function between patients in the pulsatile versus the non-pulsatile group. The evaluation is carried out via calculation of glomerular flow rate (GFR) using Cockcroft and Gault formula and 24hour urine output on a daily basis after surgery.

The secondary endpoints were other outcomes such as intraoperative and post-operative bleeding, need for blood transfusion and use of intraoperative and inpatient drugs.

### Statistics

Sample sizing was conducted via assuming a first type error of 5% and 80% power whilst considering a mean difference of outcomes of 91.58 and standard deviation (SD) of pulsatile versus non-pulsatile group of 176.08 and 106.16 regarding 24hour urine output respectively based on a preliminary sampling. A total sample size of 80 was calculated initially with two groups consisting of 40 patients in each group. Measurement data were described by mean ± SD and numerical data were described by frequency and percentage (%). Statistical differences were assessed using Pearson’s chi-square or Fisher’s exact tests on categorical variables as directed. Mann-Whitney U test independent-sample t-test was used to evaluate the differences in clinical and laboratory data. Repeated measurement tests were performed to evaluate the changes in renal function throughout ICU stay. All analyses were performed in SPSS version 23.0 and p-values less than 0.05 was considered statistically significant.

### Ethical Approval

The study was approved by the ethics committee of Shiraz University of Medical Sciences (IR.SUMS.REC.1397.108), the institutional review board, and the Iranian registry of clinical trials (IRCT 20190218042749N1, registered on 07,15,2019). It was conducted in compliance with local regulatory requirements, Good Clinical Practice (GCP), and the Declaration of Helsinki. Written informed consent was obtained from all patients or their legally authorized representatives.

## Results

Initially, a total of 80 patients were enrolled in our trial and 40 patients were included in each group of pulsatile and non-pulsatile bypass flow type. However, 16 patients included in the pulsatile group dropped out of the due to limitations in data collection and unwillingness to continue the follow-up. Figure 1 demonstrates the flowchart of the patients in our study. The demographic and baseline and clinical data of the patients in our study are provided in table 1.

**Table 1.** Patients’ demographic, intraoperative, and preliminary laboratory data

| *Variable* |  | *Total*<br>*(Mean ±SD)* | *Non-pulsatile group*<br>*(Mean ±SD)* | *Pulsatile group*<br>*(Mean ±SD)* | *P-value\** |
| --- | --- | --- | --- | --- | --- |
| *BMI* |  | *25.25 ± 4.02* | *26.04 ± 4.27* | *24.03 ± 3.31* | *0.055* |
| *Age* |  | *62.75 ± 9.97* | *63.6 ± 9.66* | *61.33 ± 10.54* | *0.383* |
| *Ejection Fraction* |  | *34 ± 6.20* | *33.27 ± 6.59* | *35.2 ± 5.41* | *0.230* |
| *Duration of*<br>*(minutes)* | *Anesthesia* | *234.14 ± 34.86* | *238.87 ± 33.34* | *226.25 ± 36.60* | *0.162* |
|  | *Perfusion* | *58.95 ± 17.83* | *60.75 ± 18.47* | *55.95 ± 16.65* | *0.302* |
|  | *Cross-clamp* | *32.84 ± 11.39* | *32.92 ± 12.86* | *32.70 ± 8.64* | *0.942* |
| *Preliminary CVP* |  | *8.68 ± 3.74* | *8.82 ± 3.91* | *8.45 ± 3.5* | *0.707* |
| *Total fluid intake* |  | *5392.34 ± 962.67* | *5300.00 ± 1064.28* | *5546.25 ± 760.42* | *0.326* |
| *Total colloid intake* |  | *632.03 ± 671.12* | *647.50 ± 825.00* | *606.25 ± 656.95* | *0.836* |
| *Urine Output on Pump* |  | *263.59 ± 237.44* | *260.75 ± 211.93* | *268.33 ± 279.69* | *0.903* |
| *Total Urine Output* |  | *626.09 ± 423.98* | *621.25 ± 433.36* | *634.16 ± 416.94* | *0.907* |
| *Ultrafiltration, UF* |  | *2539.65 ± 838.32* | *2543.24 ± 878.93* | *2533.33 ± 782.51* | *0.966* |
| *Laboratory*<br>*Data* | *BUN* | *18.26 ± 7.95* | *18.80 ± 8.47* | *17.37 ± 7.07* | *0.492* |
|  | *Cr* | *1.10 ± 0.30* | *1.14 ± 0.35* | *1.02 ± 0.18* | *0.138* |
|  | *GFR* | *69.44 ± 24.01* | *65.90 ± 21.95* | *75.34 ± 26.53* | *0.129* |
|  | *Troponin* | *1.32 ± 0.47* | *1.34 ± 0.48* | *1.29 ± 0.46* | *0.686* |

SD: Standard Deviation
BMI: Body Mass Index
CVP: Central Venous Pressure
BUN: Blood Urea Nitrogen
Cr: Serum Creatinine
GFR: Glomerular Filtration Rate
\* independent sample t-test

**Figure 1.**
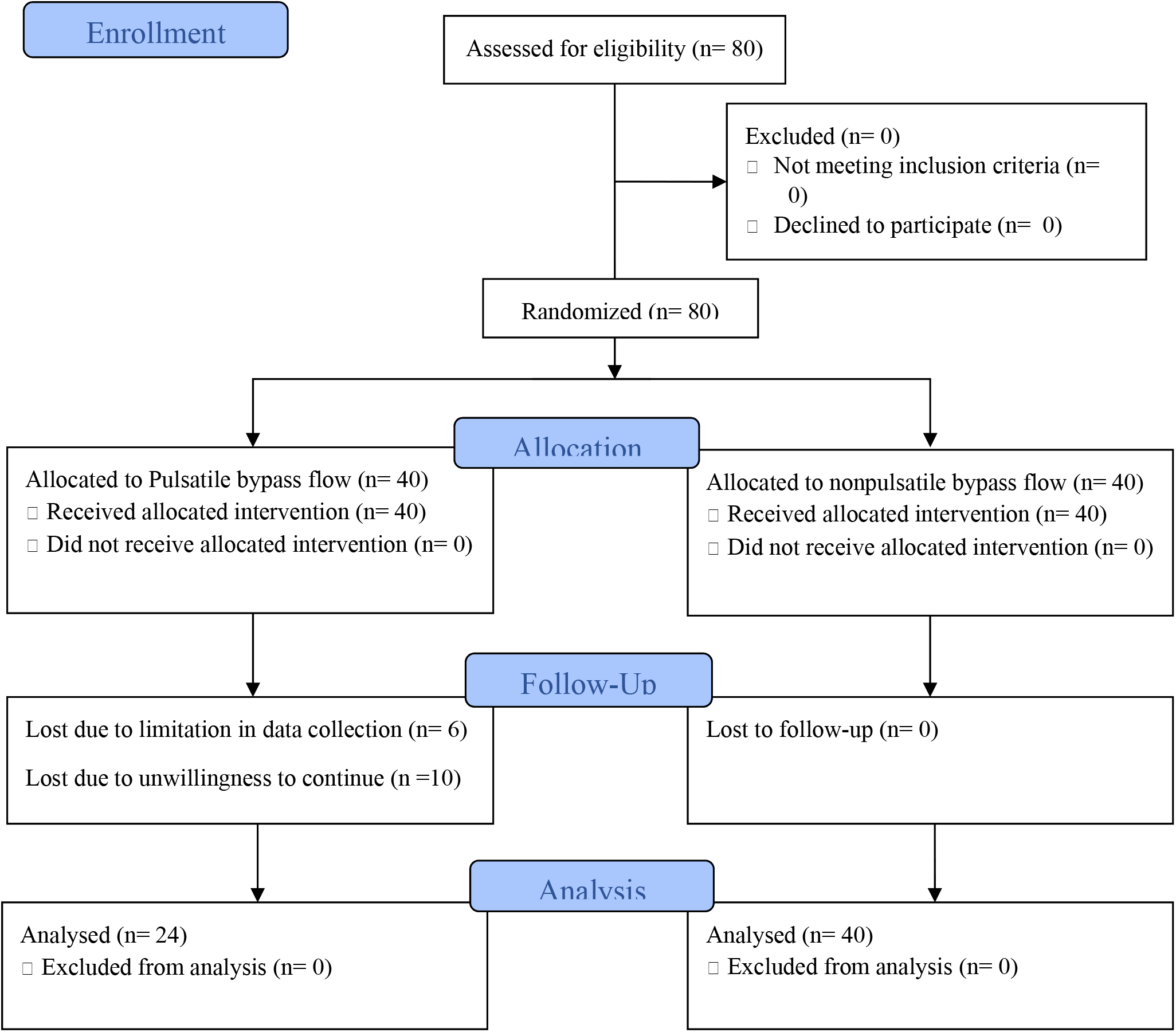
CONSORT Flow diagram of a randomized clinical trial of Pulsatile versus nonpulsatile cardiopulmonary bypass in patients undergoing coronary artery bypass graft surgery.

As illustrated in table 1, there’s no statistically significant difference in two flow type groups regarding their age, body mass index (BMI), EF, duration of phases in operation, urine output and fluid intake, and preliminary laboratory data before the intervention. The sole variable different in the groups during the operation was pack cell transfusion amount which was significantly higher in the non-pulsatile group with means of 1.72 ± 0.135 vs 0.75 ± 0.192 in the pulsatile group (P-value <0.001). Intra-aortic balloon pump was used only for 2 patients in the pulsatile and one patient in the nonpulsatile group. Differences of other intraoperative factors such as intraoperative bleeding, infusion of epinephrine and norepinephrine, and furosemide were not significant (P-value >0.05) and are available in depth in table 2.

**Table 2.** Intraoperative use of medicine, bleeding and pack cell transfusion

| *Variable*<br>*(Intraoperative)* | *Total*<br>*(Mean ±SD)* | *Non-pulsatile group*<br>*(Mean ±SD)* | *Pulsatile group*<br>*(Mean ±SD)* | *P-*<br>*value\** |
| --- | --- | --- | --- | --- |
| *Pack Cell transfusion* | *1.33 ± 0.98* | *1.72 ± 0.81* | *0.75 ± 0.94* | *<0.001* |
| *Bleeding* | *440.00 ± 156.04* | *452.50 ± 165.49* | *419.16 ± 139.74* | *0.412* |
| *Epinephrine* | *77.64 ± 31.43* | *79.67 ± 31.40* | *73.23 ± 32.01* | *0.490* |
| *Norepinephrine* | *84.00 ± 42.60* | *71.25 ± 34.82* | *98.57 ± 48.53* | *0.228* |
| *Furosemide* | *20.00 ± 0.00* | *20.00 ± 0.00* | *20.00 ± 0.00* | *N/A* |

*SD: Standard Deviation*
*N/A: Not Applicable*
*\* Independent samples t-test*

A sum of 24 of 40 patients in the non-pulsatile group had an ICU length of stay of more than two days versus 16 that had two. Similarly, 13 patients of the pulsatile group had a two-day ICU length of stay while 11 stayed in the ICU ward for more than two days.

The patients’ post-operative features during their ICU stay, along with laboratory data changes and administered drugs were compared among the two groups in our study. Regarding our main objective through this study, renal function of the patients between the pulsatile and nonpulsatile flow type groups were compared before operation and during their ICU stay using repeated measurements test.

The changes in GFR in this timeframe was significant in all patients (P-Value <0.001). However, when comparing the changes in GFR between two groups, there was no statistically significant difference amongst these two groups (P-Value =0.970).

The same rule applied for changes in serum creatinine levels and BUN, both of which had significant changes while compared without considering the two different groups (P-value <0.001). However, these changes did not differ significantly between the two groups with P-values equal to 0.496 and 0.916 in the pulsatile and nonpulsatile group, respectively. To furtherly compare these two groups, repeated measurements test was performed on each group individually and the changes in GFR, BUN and serum creatinine levels were statistically significant (P-Value <0.001). At the same time, differences in these laboratory data were also compared before and in each of the three days of ICU stay between the two groups using independent-samples t-test and the differences in means were not statistically significant. 24-hour Urine output was also recorded in 3 days of ICU admission but the changes in this factor was as same as the laboratory data recorded in these patients meaning even though the changes in 24-hour urine output through days of admission was statistically significant, these changes were not statistically significant when compared across the two groups of pulsatile and nonpulsatile group. The changes in our primary targets are illustrated in figure 2.

**Figure 2.**
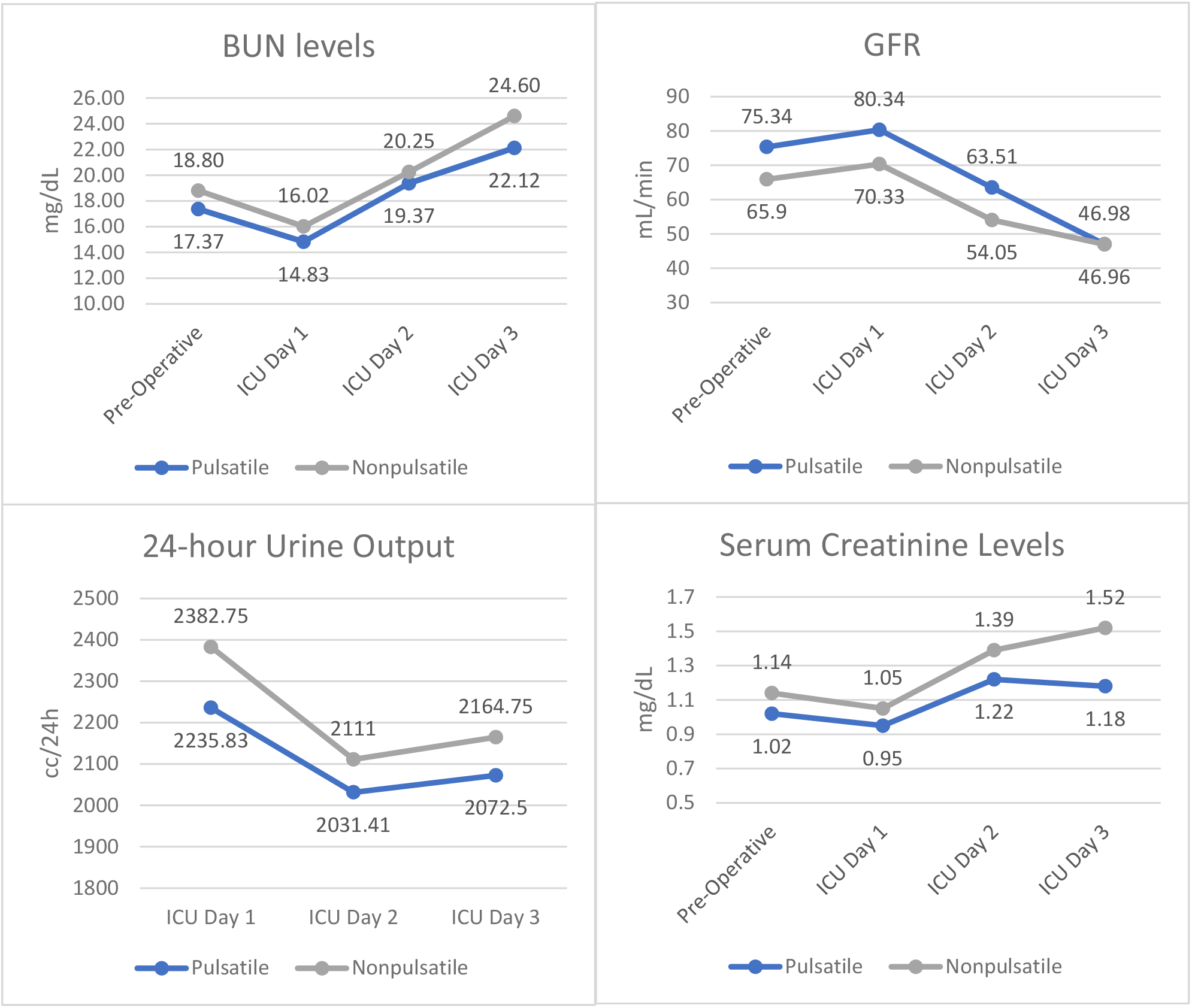
Changes in renal function target variables in pulsatile versus nonpulsatile flow type groups

Regarding the duration of intubation in the ICU, the pulsatile flow group showed a statistically significant lower duration of intubation of 12.87 ± 9.85 hours versus 18.30 ± 4.64 in the nonpulsatile group (P-Value =0.014). Regarding the secondary objectives of this study, bleeding volume, pack cell transfusion, CVP, central nervous system complications, cardiac arrythmia, and epinephrine, norepinephrine, and furosemide administration was also recorded and compared between the groups. None of the patients experiences any central nervous system complications or cardiac arrythmia in their first three days of ICU stay. Comparing other measurements in the first three days ICU admission, bleeding was the only variable that had a statistically significant difference between the two groups. Patients in the nonpulsatile group had less bleeding than the patients in the pulsatile groups with a bleeding volume of 580.15 ± 477.91cc versus 900.00 ± 511.39cc respectively (P-Value =0.014). The other factors measured in this study did not have any statistically significant changes regarding either repeated measures in consecutive days of ICU admission or in between the two groups.

## Discussion

Despite numerous and extensive studies conducted on different flow types during CPB, the controversy remains since these studies have failed to prove a superiority of each of the discussed flow types(14). Our study has demonstrated that the choice of different flow types does not show any impact on the patients’ outcome regarding renal insufficiency. All patients who underwent cardiopulmonary bypass showed an increase in serum creatinine levels leading to a decrease in GFR during their ICU stay and difference was observed between the patients with pulsatile and nonpulsatile flow type. The only measurements significantly different between the two groups were intraoperative blood transfusion, bleeding in the ICU ward and intubation duration. The patients in the pulsatile group had an average of about 5.5 hours less intubation time than the patients in the nonpulsatile group. Also, the patients in the pulsatile group had a significantly less blood transfusion intraoperatively. This in part could be noted since blood transfusions come with various risks and complications. However, the patients with pulsatile blood flow during surgery had a higher bleeding volume in their first day of ICU stay.

As well may some studies prove pulsatile perfusion leading to less microcirculatory complications, it appears further investigation shows there’s little to no difference regarding the big picture being the clinical outcome of such patients(15-17).Poswal et. al. have illustrated that even though patients had a different creatinine clearance rate post operatively, there was not a statistically significant difference between the groups at discharge(18). Adademir et. al, illustrated the use of nonpulsatile flow leads to a higher concentration of Urinary neutrophil gelatinase-associated lipocalin and interleukin-18, both markers of renal injury(19). Presta et. al, have shown that even though myocardial revascularization leads to a decrease in GFR and hence the renal function in all patients, pulsatile CPB leads to a less decrease and therefore preserves the renal function better than nonpulsatile flow(20). This study was conducted as an extension to the one conducted by Onorati et. al, in order to furtherly investigate the role of patients’ age since ageing impacts the kidney and its function both anatomically and physiologically(21, 22).Our study had a relatively younger population involved and thus may show a broader view on flow types than when involving a more senile population. Hornick et. al have illustrated pulsatile blood flow may be beneficiary in certain identifiable high risk patient groups (23). This could prove that the statistics reported by other studies may be due to the patient selection rather than the differences in blood flow types in CPB. Sink et. al have also shown that even though differences in these two flow types has not been statistically significant, these differences must be explained via differences in other factors not evaluated in their study(24). Similarly in a meta-analysis by Sievert et. al, selected high risk patients for renal failure are the main subjects whom could benefit more from the choice of pulsatile blood flow(25).

## Limitations

The only limitation of this study was exclusion of patients due to shortcomings in data gathering and their unwillingness to continue their participation in this study.

## Conclusion

This study has illustrated that there is no statistically significant difference regarding renal function between patients with pulsatile or non-pulsatile blood flow in cardiopulmonary bypass. Our study has had a broader age range and a relatively acceptable number of age range compared with similar studies.

## Data Availability

The data that support the findings of this study are available on request from the corresponding author, M. Najafi. The data are not publicly available due to ethical committee's decision regarding privacy of patient data.

## Notes

### Competing Interest Statement

The authors have declared no competing interest.

### Clinical Trial

IRCT20190218042749N1

### Funding Statement

This research received no specific grant from any funding agency in the public, commercial, or not-for-profit sectors.

### Author Declarations

This study was approved by shiraz university of medical sciences ethics committee. IR.SUMS.REC.1397.108

